# The online delivery of exercise oncology classes supported with health coaching: A pilot randomized controlled trial

**DOI:** 10.1101/2021.11.29.21266169

**Authors:** Maximilian Eisele, Rosie Twomey, Andrew J. Pohl, Meghan H. McDonough, Margaret L. McNeely, Manuel Ester, Julia T. Daun, S. Nicole Culos-Reed

**Author notes:** **Corresponding Author:** Maximilian Eisele MSc.

## Abstract

**Purpose:** The primary objective was to investigate the feasibility of a synchronous, online delivered, group-based, supervised, exercise oncology maintenance program supported with health coaching.

**Methods:** All participants had previously completed a 12-week group-based exercise study. In the current study, participants were randomized to a 12-week exercise oncology maintenance class with or without health coaching. The primary outcome was feasibility, assessed as intervention attendance, safety and fidelity, study recruitment, attrition and outcome assessment completion. Additionally, semi-structured interviews at the end of the intervention provided participants’ perspectives on intervention feasibility.

**Results:** Forty participants (n_8WK_=25; n_12WK_=15) enrolled in the study. Feasibility was confirmed for recruitment rate (42.6%), attrition rate (2.5%), safety (no adverse events), health coaching attendance (97%), health coaching fidelity (96.7%), class attendance (91.2%), class fidelity (92.6%), and assessment completion (questionnaire=98.8%; physical functioning=97.5%). Based on the qualitative feedback, feasibility was facilitated by the convenience, while the diminished ability to connect with other participants online was a drawback compared to in-person delivery.

**Conclusion:** The synchronous online delivery of an exercise oncology maintenance class, the additional health coaching support, and the tools used to measure the intervention effectiveness were feasible for individuals living with and beyond cancer.

## Introduction

The COVID-19 pandemic and associated physical distancing measures have limited access to fitness facilities and sporting activities, increased sedentary behaviour, loneliness, and reduced physical activity (PA) levels^1^. These issues are exacerbated for individuals living with or beyond cancer^2^, who may be dealing with negative side effects of cancer and cancer treatment and may be immunocompromised^3^. Indeed, these individuals face more than a two-fold risk of contracting COVID-19 and are at increased risk for more severe symptom progression^4^. PA reduces the impact of cancer-related side effects such as fatigue^5^, cachexia and cardiorespiratory deconditioning^6^, so necessary physical distancing measures may be more detrimental in this population^3^.

While PA is beneficial for individuals living with and beyond cancer^7,8^ and evidence has supported the development of cancer exercise guidelines^9^, the proportion of individuals meeting these guidelines was low even before the pandemic^10^. Common barriers to being physically active include lack of time, proximity to exercise facilities or accessibility of cancer-specific exercise programs, and treatment-related symptoms^11^. These barriers, and new barriers imposed by physical distancing measures (e.g., facilities being closed), can be addressed by providing home-based exercise oncology programs.

To date, most home-based exercise oncology interventions have been unsupervised and reported lower adherence rates compared to in-person supervised settings^12-14^. The synchronous delivery of online exercise programming by a trained professional may address barriers to PA maintenance (e.g., accessibility to facilities and time constraints). However, to our knowledge, the feasibility of providing synchronous online group-based exercise programming for individuals living with and beyond cancer, with virtual supervision by a qualified exercise professional has yet to be examined. Synchronous online supervised exercise interventions in other populations with chronic disease have shown promising results in terms of feasibility and preliminary effectiveness^15,16^. To support online exercise interventions and the long-term maintenance of increased PA, additional support may be beneficial, and health coaching (HC) provides one option. Health coaching is a behaviour change tool that is participant-centred and built on a coach-participant relationship^17^. It includes participant-determined goals, a self-discovery process to find solutions, participant accountability, and health education^17^. Preliminary evidence suggests HC may increase QoL, mental well-being^18^, PA levels^19^, as well as maintenance of PA levels^20,21^. Given these potential benefits, the primary objective of this pilot study was to assess the feasibility of a synchronous online group-based supervised exercise oncology maintenance program with additional HC support. We hypothesized that an online supervised group-based exercise oncology program supported with HC would be feasible, as measured by a class attendance rate of ≥ 70%, a HC completion rate of ≥ 80%, and an assessment completion rate of ≥ 70%. Recruitment rate, safety, and fidelity of the class and HC were also evaluated. The qualitative inquiry aimed to understand the participants perceptions on the feasibility of completing assessments, participating in synchronous online exercise classes and weekly HC calls, as well as understand how the intervention may have prepared them to be successful in maintaining being active.

## Methods

### Study Design

The study was a pilot randomized controlled trial (RCT) with two parallel intervention arms. An embedded mixed methods study design was used, guided by a pragmatic philosophy, which aligned with addressing practical concerns of feasibility^22^. The study was approved by the Health Research Ethics Board of Alberta – Cancer Committee (HREBA.CC-19-0206) and retrospectively registered as a clinical trial (NCT04751305). The retrospective registration was due to the rapidly evolving COVID-19 situation that precipitated the rapid switch to online programming.

### Study Setting

All components of the study were performed in an online environment through an end-to-end encrypted version of the Zoom videoconferencing application (Zoom video communications; San Jose, CA) or the survey monkey platform (Momentive Inc.; San Mateo, CA) for patient-reported outcomes. Two waves were conducted – as a result of initial COVID-19 delays, the first wave was 8 weeks long, and the second wave was 12 weeks long, as intended. Recruitment, trial commencement, and follow-up for the first wave occurred in May, end of May, and end of July 2020, respectively. Recruitment, trial commencement, and follow-up for the second wave occurred in August, the beginning of September, and the beginning of December 2020, respectively. The only change made to the study methods after the trial commenced was to add an activity tracker usage questionnaire to the post-assessment for the 12-week participants.

### Participants

The inclusion criteria were 1) 18 years or older; 2) completed the baseline Alberta Cancer Exercise (ACE) program (12-week introductory group-based exercise class, delivered in-person for the first group in the current study; and delivered online for the second group); 3) access and familiarity with a computer, laptop, or tablet with a video camera capable of running Zoom video conferencing software; 4) an internet connection strong enough to support a live video broadcast; and 5) provided written informed consent in English. Participants were also screened for exercise readiness by a clinical exercise physiologist (CEP) with the PA Readiness Questionnaire tool. Eligible participants completed the baseline assessments and were randomized with a 1:1 allocation ratio to either the HC intervention or the non-HC intervention through an online random sequence generator ^23^. The first author enrolled participants, generated the random allocation sequence, and assigned participants to the intervention groups. The class instructors and physical functioning assessors were blinded to intervention allocation (HC or or non-HC). All participants that completed the intervention were invited to participate in semi-structured interviews. The goal was to interview half of each of the intervention (HC and non-HC) groups to gather feedback on the intervention components from a range of participants. Interviews were audio-recorded over the Zoom application.

#### Online Exercise Oncology Maintenance Program

All participants received the online group-based exercise oncology maintenance program, which consisted of two components – online exercise classes and additional resources to support additional home-based exercise engagement. An in-depth description of the intervention, including the behaviour change support, can be found in supplementary file 1. The classes were instructed by a CEP through Zoom video conferencing and were multimodal, including strength, cardiovascular fitness, balance, and flexibility exercises. The CEP was assisted by a moderator during class, whose responsibilities were to ensure safety and address technical problems with the Zoom application ^24^. After each class, the moderator also facilitated a post-class discussion intended to foster social support, evoke thoughts about an active lifestyle (e.g., by posing discussion questions), and offer the opportunity for questions. The classes were offered twice a week for the first two weeks of the program and then reduced to once a week for the remainder of the study to facilitate participants being active independently for the remainder of the week. Once tapering occurred, all participants received a PDF-based at-home exercise program, which included six different circuits with three different intensity options, providing the participants with the opportunity to tailor exercises to meet their needs.

#### Health Coaching Intervention

Participants randomized to the HC intervention received weekly individual HC calls. HC was structured based on Wolever et al.^17^, where HC is participant-centred, built on a coach-participant relationship, and includes participant-determined goals, a self-discovery process to find solutions, patient accountability, and education. A day before each HC call, the participants completed a survey on fatigue, QoL, stress, loneliness, and social support, enabling tailoring of the HC call to address individual needs.

Educational topics within the HC calls included: Goal Setting, Monitoring Behaviour, Barrier Management, Social Support, Stress Management, Adapting the Program, Self-compassion*, Sleep & Nutrition*, Reflection*, Health Media*, Remote Resources, and Maintaining Motivation. The order of the educational topics could be adjusted to meet participant needs. Each HC call was structured, starting with a reflection on the previous week, a conversation about the educational topic, and finishing with an action plan for the upcoming week. Based on participant interest, a summary sheet of the educational topic was sent to the individual. At the halfway point of the intervention, the participant provided feedback on the HC calls by setting time aside at the end of the HC call. The health coaches were graduate students trained in behaviour change strategies, exercise oncology, had extensive experience with the larger ACE program, and completed at least 30 hours of HC training (mock interviews, motivational interviewing techniques, and reviewing the HC literature). The weekly HC calls were held via Zoom at a convenient time for the participant, and the length of each call was dependent on the participant’s needs. No restrictions on receiving additional counselling or coaching from outside sources were made. The behaviour change tools used can be found in Supplementary file 1.

#### Primary Outcome: Feasibility

Recruitment was calculated as the percent of those that participated in the study from those eligible (with no pre-determined level). Safety and adverse event reporting followed standardized guidelines^25^ and were tracked by the moderator. After completion of the intervention, all participants were invited to participate in semi-structured interviews, which focused on their perceptions and satisfaction with the online classes, the HC intervention, the assessments, and their perceptions of safety. Recruitment, safety and adverse event reporting and fidelity did not have pre-determined criteria (levels) of feasibility due to limited previous literature of recruiting and administering an online exercise intervention to individuals living with and beyond cancer. Feasibility measures with pre-determined levels included online exercise and HC attendance, as well as completion of assessments.

##### Online Exercise Classes

The criteria for establishing the feasibility of the scheduled exercise classes was set at 70% attendance, based on previous findings in an online synchronous setting^16^. Assessment of intervention fidelity (i.e., delivery of content and timing as intended) was performed using a structured fidelity checklist that was completed by the moderator during each class. Fidelity was reported as the overall percentage of items on the checklist adhered to across all classes. Participants were asked about barriers and facilitators to exercise class attendance during interviews.

##### Health Coaching

The feasibility criteria for the HC attendance was set at 80%, based on HC completion rates reported in exercise oncology^20^. The fidelity of the HC delivery was assessed by randomly recording two HC calls for each participant. HC recordings were assessed by independent evaluators with HC training, comparing the HC protocol to the recorded HC call. HC call fidelity was reported as the overall percentage of items adhered to on the checklist across the recorded HC sessions. Additionally, participants were asked in the interview about their preference regarding the HC call structure in terms of length, frequency, and general delivery.

##### Assessments

The feasibility cut-off for the completion rate of the physical functioning assessments, the objective PA data, and the questionnaires was set at 70%^26^. Participants’ perspectives of these feasibility aspects of the collected data were also explored in the interview.

#### Exploratory Outcomes

Outcomes assessing the potential impact of the intervention are not included in the current manuscript but can be found in Supplementary file 4. Individual-level assessments included online physical functioning assessments, PA level assessments (objective and subjective), and patient-reported outcomes. All measures occurred pre- and post-intervention, except for objective PA levels, which were reported continuously throughout the intervention via a wearable device. For details on measures and the exploratory results, refer to Supplementary file 3 and 4.

#### Analysis

##### Quantitative Data Analysis

All data were analyzed using SPSS statistics (v26, IBM). For continuous data, normality was assessed by inspecting histograms, box plots, QQ-plots, and the Shapiro-Wilk test of normality. Descriptive statistics were reported as means and standard deviations (SD) for normally distributed data and medians and interquartile ranges (IQR) for non-normally distributed data. Categorical outcomes were reported as frequencies and percentages.

##### Qualitative Data analysis

Audio recordings were transcribed verbatim, and data were managed in NVivo12 (QSR International 2019; Burlington, MA). Data analysis used qualitative description^27^ and illustrative analysis^22^. Throughout the analysis, the first author stayed close to the participant responses and simply described the content by assigning quotes to shared beliefs. The qualitative results were compared with and interpreted in light of the quantitative results to illustrate and provide context and elaboration on the feasibility of the intervention. The quality of the qualitative portion of the study was assessed based on its credibility, dependability, transferability, and confirmability^28^. Credibility was enhanced through prolonged engagement (the interviewer moderated each class and health coached some of the participants) and methodological triangulation of the qualitative and quantitative data. To enhance transferability, we strove for thick description (to the extent qualitative description allowed) to allow readers to form their own interpretation of the applicability of the findings to other situations. Dependability and confirmability were enhanced by a dependability and confirmability audit respectively, with the senior author being the auditor.

## Results

### Participant Demographics

The study included 40 participants (25 participants in the 8-week wave and 15 in the 12-week wave). The reasons for exclusion are outlined in Figure 1. The mean participant age was 56 ± 9 years and most study participants self-identified as being of European ancestry (82.5%) and female (92.5%). The most common cancer diagnosis was breast cancer (70.0%) and more than half of the participants were on active treatment during the intervention (55.0%). The most common treatment was surgery (87.5%), followed by chemotherapy (75.0%) and radiation (62.5%). Half of the participants had previous experience with an activity tracker (50.0%). A full description of demographics can be found in table 1. Approximately half (19/39, 48.7%) of the eligible participants participated in the post-intervention interviews. Of these participants, 10 were from the non-HC group (10/20; 50%) and 9 were from the HC group (9/19; 47.4%).

**Table 1.** Participant demographics for the 8- and 12-week interventions

| Group | Wave 1 (8WK) |  | Wave 2 (12WK) |  | Total |
| --- | --- | --- | --- | --- | --- |
| | HC<br>(n = 12)<br>(Mean ± SD<br>or<br>n (%)) | Control<br>(n = 13)<br>(Mean ± SD or<br>n (%)) | HC<br>(n = 7)<br>(Mean ± SD<br>or<br>n (%)) | Control<br>(n = 8)<br>(Mean ± SD<br>or<br>n (%)) | (n = 40)<br>(Mean ± SD or<br>n (%)) |
| Age (in years) | 59 ± 10 | 54 ± 10 | 54 ± 6 | 56 ± 8 | 56 ± 9 |
| Sex |  |  |  |  |  |
| Female | 11 (91.7) | 12 (92.3) | 7 (100) | 7 (87.5) | 37 (92.5) |
| Male | 1 (8.3) | 1 (7.7) | 0 (0) | 1 (12.5) | 3 (7.5) |
| Education status |  |  |  |  |  |
| High School or less | 1 (8.3) | 0 (0) | 0 (0) | 2 (25) | 3 (7.5) |
| Some University or more | 11 (91.7) | 13 (100.0) | 7 (100) | 6 (75) | 37 (92.5) |
| Employment Status |  |  |  |  |  |
| Working – Part/Full Time | 2 (16.7) | 3 (23.1) | 3 (42.9) | 2 (25) | 10 (25) |
| Not Working – Retired,<br>Disability, Unemployed | 10 (83.3) | 10 (76.9) | 4 (57.1) | 6 (75) | 30 (75) |
| Marital status |  |  |  |  |  |
| Married | 12 (100) | 6 (46.2) | 7 (100) | 7 (87.5) | 32 (80) |
| Common Law | 0 (0) | 1 (7.7) | 0 (0) | 0 (0) | 1 (2.5) |
| Widowed | 0 (0) | 2 (15.4) | 0 (0) | 0 (0) | 2 (5) |
| Divorced | 0 (0) | 4 (30.8) | 0 (0) | 0 (0) | 5 (12.5) |
| Ethnicity |  |  |  |  |  |
| European | 10 (83.3) | 12 (92.3) | 4 (57.1) | 7 (87.5) | 33 (82.5) |
| Indigenous | 0 (0) | 0 (0) | 0 (0) | 1 (12.5) | 1 (2.5) |
| East and South Asian | 2 (16.7) | 1 (7.7) | 1 (14.3) | 1 (12.5) | 5 (12.5) |
| Latin/Central and South<br>American | 0 (0) | 0 (0) | 1 (14.3) | 0 (0) | 1 (2.5) |
| African | 0 (0) | 0 (0) | 1 (14.3) | 0 (0) | 1 (2.5) |
| Mixed | 1 (8.3) | 0 (0) | 0 (0) | 0 (0) | 1 (2.5) |
| Cancer type |  |  |  |  |  |
| Breast | 6 (50) | 10 (76.9) | 6 (85.7) | 6 (75) | 28 (70) |
| Lung | 0 (0) | 1 (7.7) | 0 (0) | 0 (0) | 1 (2.5) |
| Vascular | 0 (0) | 2 (15.4) | 1 (14.3) | 0 (0) | 3 (7.5) |
| Gynecological | 4 (33.3) | 1 (7.7) | 0 (0) | 1 (12.5) | 6 (15) |
| Genitourinary | 1 (8.3) | 0 (0) | 0 (0) | 1 (12.5) | 2 (5) |
| Head and Neck | 1 (8.3) | 0 (0) | 0 (0) | 1 (12.5) | 2 (5) |
| Skin | 1 (8.3) | 0 (0) | 1 (14.3) | 2 (25) | 4 (10) |
| Active cancer treatment | 5 (41.7) | 7 (53.8) | 3 (42.9) | 7 (87.5) | 22 (55) |
| Using an activity tracker | 4 (33.3) | 9 (69.2) | 3 (42.9) | 4 (50) | 20 (50) |

**Figure 1.**
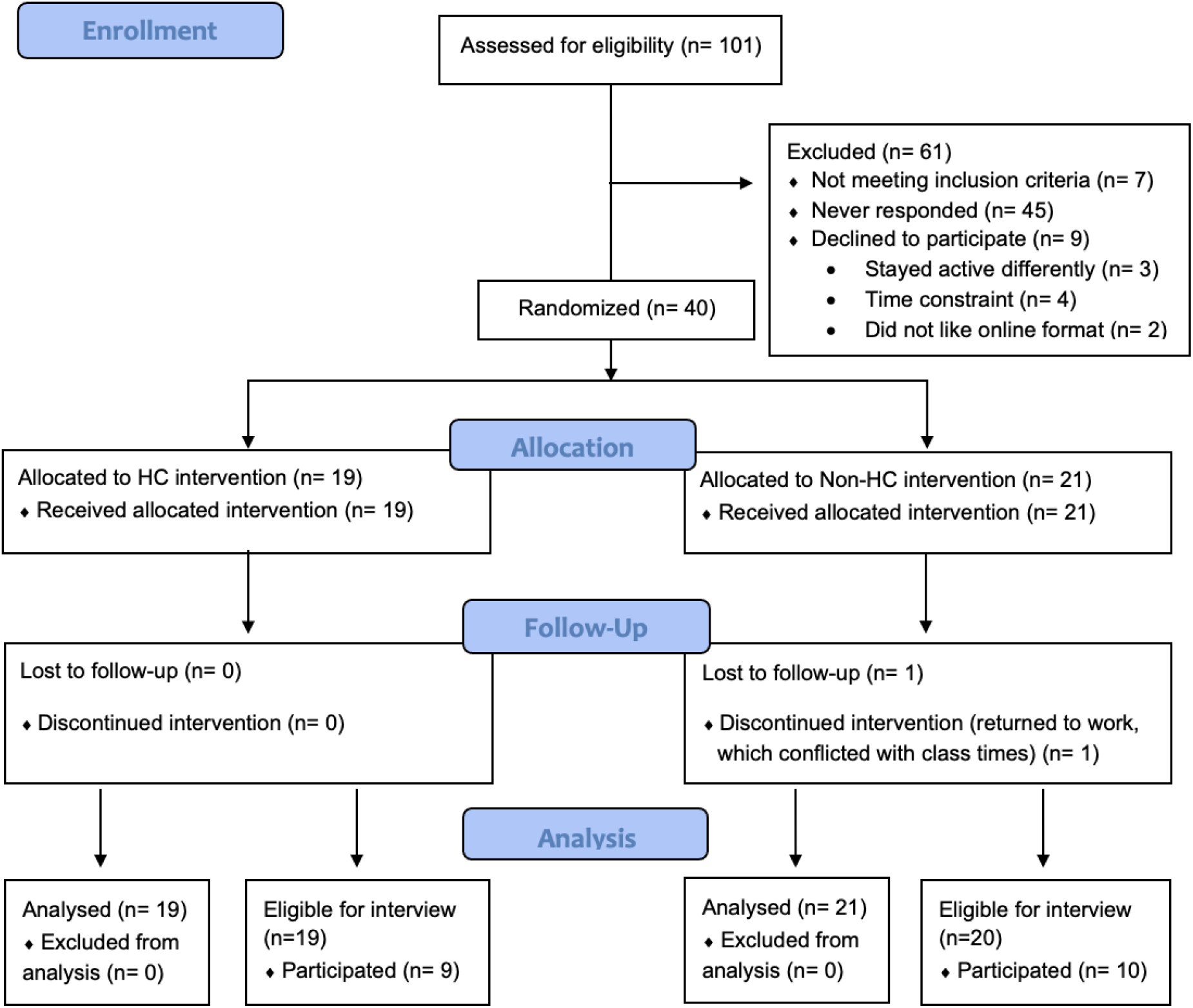
Flow of Participant Recruitment, Randomization, and Attrition

### Feasibility

The overall study recruitment rate was 42.6% (Figure 2). The attrition rate of the study was 2.5%, with one participant dropping out after 4 weeks due to returning to work. No adverse events were reported. A summary of the quantitative feasibility results can be found in table 2.

**Table 2.** Feasibility and safety results of the synchronous online exercise maintenance study for individuals living with and beyond cancer

| Feasibility & Safety Measure | Frequency n (%) |
| --- | --- |
| Recruitment Rate | 40/94 (42.6) |
| Qualitative Interview Recruitment Rate | 19/39 (48.7) |
| Attrition Rate | 1/40 (2.5) |
| Adverse Events | 0 (0) |
| Completion Rate |  |
| Questionnaires | 79/80 (98.8)* |
| Objective PA Assessment | 1489/1785 (83.4)* |
| Physical Functioning Assessment | 78/80 (97.5)* |
| Attendance Rate |  |
| Online Exercise Class | 401/441 (91.2)* |
| Post Online Exercise Class | 390/441 (88.4) |
| Health Coaching | 163/168 (97.0)<sup>o</sup> |
| Fidelity |  |
| Online Exercise Class | 376/406 (92.6) |
| Health Coaching Call | 294/304 (96.7) |

\*Feasibility cut-off set a priori: $\geq 70\%$
<sup>o</sup>Feasibility cut-off set a priori: $\geq 80\%$

#### Online Exercise Classes

Class attendance was 91.2%, with a 13.5% higher attendance rate for the HC (94.5%) vs the non-HC group (81%). A similar trend was observed in the post-class discussion, with an overall attendance of 88.4%, a HC group attendance of 91.7%, and a non-HC attendance rate of 78.5%. Facilitators of exercise program attendance reported during the interviews included convenience (n=13/19), reduced concern about physical appearance (n=4/19), and less exposure to pathogens, which was important for this immunocompromised population (n=3/19). Participants mentioned reduced travel time to the exercise facility, being in the comfort of home, and not being location-bound contributed to convenience.

> *But you know at 5:00 o’clock in the city, … I wouldn’t be driving over to the University and back at 6. Like that would be, you’d be right in traffic at that time. So that’s pretty convenient just to turn on your iPad and get going*. (female, non-HC)

Barriers to online programing attendance included the equipment availability, the ability to tailor exercises to individuals, and the reduced chances to connect with other individuals living with and beyond cancer as compared to in-person programs:

> *I guess that’s probably the disadvantage of the virtual, we didn’t really get to meet each other on a kind of … a more social level before and after class that we normally would’ve … had we been in-person. And I think … as much as the virtual was a good backup, in-person would be great. … Largely because I think it just gives everybody the support because we’ve all gone through our own little personal hell*. (female, non-HC)

Fidelity of the exercise class delivery was high, with 92.6% of the exercise classes following the protocol as intended, with technical interruptions (4.4%), participants arriving more than five minutes late to class (2.5%), and the class instructor altering the sequence of the exercises (0.5%), reported as protocol deviations. In interviews, participants mentioned their preferred structure would be classes at least twice a week (n=18/19) for an hour duration each session (n=19/19). There were no adverse events during any of the classes, and all interviewed participants felt safe during classes (n=19/19). Program components that enhanced participants’ perceptions of safety included receiving the exercise program beforehand, which allowed them to look up a video of each exercise, and having two exercise oncology fitness professionals present during each class:

> *Yes, but being on camera I think didn’t make me worry that I was doing something unsafe because I knew that the instructor and the person that was watching … would correct you if there was something that was unsafe*. (female, HC)

#### Health Coaching

The completion rate for the HC calls was 97.0% (8WK: 6.8/7 sessions completed; 12WK: 11.6/12 sessions completed). Reasons for missing a call included vacation, having a migraine, contracting COVID-19, and being stuck at work. The mean call length was 34.2± 13.2 minutes, with a range from 20.2 ± 4.0 to 52.3 ± 11.2 minutes. The fidelity of the HC sessions being delivered as intended was 96.7%. Additionally, most participants preferred receiving HC calls once per week, as that schedule gave sufficient time to implement the weekly goals while keeping them accountable.

Aspects of HC as a tool to support exercise engagement included fostering connection with the health coach, providing tailored educational topics that addressed individual needs, and having an active listener to keep one accountable and motivated.

> *Having somebody there who is not your husband, your spouse, your child, your family member who is there and who is committed to your well-being as much as you are. And who will give you some advice, who will provide a listening ear, who will give you some encouragement, and who will put things* … *into perspective. … So, it was valuable, like it was really good, I will definitely highly recommend having that*. (female, HC)

#### Assessments

The questionnaire completion rate was 98.8% and within completed questionnaires, 98.4% of questions were answered. Participants did not find questionnaires to be burdensome, appreciated being able to return to complete them at a later time via the online system, but noted that it would have been helpful to be able to add context or indicate if a question was not applicable:

> *I’ve been out of treatment for a year and had a really good checkup yesterday by the way and so yeah so some of those questions weren’t necessarily as relevant to me*. (female, non-HC)

The completion rate of the physical functioning assessment was 97.5%, with two participants not completing the final assessment. There were no adverse events or safety concerns. Both participants who did not complete the physical functioning assessment were in the non-HC group: one discontinued the study after 4 weeks, and the other was injured (unrelated to the study) in the last week of the study. A facilitator for assessment completion commonly mentioned by participants was their interest in seeing results:

> *I found it quite easy and it was kind of neat I found I … maintained most of almost all of my levels and I increased my cardio so that was good*. (female, non-HC)

Despite the feasibility of the online physical functioning assessments, some participants did not feel particularly confident in the results. This doubt was especially apparent for the hamstring flexibility measure:

> *Yeah, that’s a little difficult you know like (laughs) only because I was like I’m using a ruler to try to measure you know like how far I can stretch […] so I’m not sure if it was 100% accurate*. (female, HC)

Of the participants with available data (n=36), the trackers were worn ≥10h per day 83.4% of the time, and they were worn on average 5.84 ± 1.87 days per week. Between intervention groups, the non-HC group wore the tracker for 84.7% and HC group for 82.1% of the possible days in the study. In both waves, the wear time was over the 70.0% feasibility threshold, except for the last week of the 8-week wave (68.9%). No data was obtained from 4 participants: one participant dropped out before receiving the tracker, one participant did not sign the updated consent form regarding Garmin data storage, and two participants synchronized their tracker with a private account instead of the research account. One participant received the tracker a week late due to postal delays and therefore did not have data for the first week. In general, a trend that was visible in both waves was that the wear time was lower in the first and last week of wearing the tracker, as seen in Supplementary file 4.

## Discussion

A synchronous online group-based supervised exercise maintenance program is safe and feasible for individuals living with and beyond cancer. Adding health coaching to this program is also feasible, based on quantitative data (attendance and fidelity above pre-specified thresholds) and participant feedback. Finally, this pilot RCT shows a full RCT will be feasible because recruitment was in line with our in-person exercise maintenance programs, and outcome assessments could also be completed online. Improved practices for the objective measurement of physical activity (PA) levels were learned during this study, and these can be adopted in future trials.

In the move from in-person to synchronous, group-based online exercise programming, safety was a key consideration. To ensure safety, our protocols included extensive staff training on verbal and visual cueing, and a moderator (in addition to the instructor) was present throughout classes, whose primary responsibility was to ensure participant safety and well-being. Participants were required to always remain on camera on Zoom^24^, and an emergency response plan was also in place. No adverse events occurred during this study, and these safety protocols should be implemented in future online programs, while exploring where measures could be loosened to increase accessibility. The online exercise classes were delivered as intended with a high fidelity rate, and attendance was higher compared to previous synchronous online classes^15,16^. Our high attendance may be because classes took place during COVID-19 pandemic, where fewer in-person options were open, and people may have been and motivated to attend online classes. One consideration with online exercise classes is the risk of technical interruptions, but these only occurred in 4.4% of the classes, much lower compared to the 25.0% of sessions interrupted in a study by Tomlinson et al.^16^. This may be due to the increased emphasis participants placed on having a good internet connection during the pandemic, and thus the growing potential for synchronous delivery of online exercise programs^24^. However, as most of our participants were based in urban areas, it is possible that internet bandwidth may be a larger barrier to participating for individuals in rural or remote locations. The delivery of synchronous group-based supervised exercise classes to individuals living with and beyond cancer addresses important access issues both during and post-pandemic, particularly for immunocompromised participants or those who are unable to travel while still allowing them access to resources to support their health and wellness^29^. Similar to earlier work^16^, the removal of travel time was the most frequently mentioned factor that contributed to the convenience of the home-based program. Our online program also eliminated another common barrier to PA; the distance to exercise facilities^11^.

A high proportion of health coaching calls were completed (97%), and overall, participants were positive about taking part in this aspect of the study. Our finding on the feasibility of health coaching supports the small number of previous studies in exercise oncology^20,21,30^. The mean HC call length (34.2 minutes) aligns with a recent study in individuals living with and beyond cancer that recorded a median call length of 31.5 minutes^20^. Although education is defined as one of the key pillars of HC^17^, other studies in exercise oncology did not include this component within their HC, and this generally resulted in shorter call durations (18-24 minutes)^31,32^. The duration of the HC calls has both pragmatic and economic implications for sustainable implementation^33^ and aiming for a 30-minute call length that includes the key HC components was feasible in our setting. The coach-participant relationship, accountability, and the tailored educational components were highlighted by participants as important components of the HC sessions, fostering PA attendance and a sense of well-being. It is important to note that these components are part of the definition of HC^17^, indicating the HC intervention was both delivered (fidelity of 96.7%) and received by participants as intended. It is critical that future studies structure HC based on Wolever et al.’s^17^ definition and report on both effectiveness and costs because one-on-one HC is relatively resource-intensive. Future work could tailor HC to those ‘most in need’of additional behaviour support for and maintenance of PA levels (i.e., those with low attendance, those without other sources of social support).

Several other components of this study suggested that the intervention could be tested in larger trials. Firstly, the recruitment rate of 42.6% for the online exercise maintenance study was comparable to the recruitment for our previous in-person exercise maintenance program (41.3%). Other studies with remote delivery of exercise supported with HC had recruitment rates ranging from 35.9% ^20^ to 70.2% ^21^. However, recruitment may have been lower than we might expect in a future trial because the community was dealing with additional personal stressors during the COVID-19 pandemic, especially those navigating cancer treatment^34^. Future research could explore preferences for online versus in-person delivery once in-person exercise programming resumes. Online programs are beneficial for people who live in remote rural areas, and some individuals may prefer to continue in virtual programs. Assessments of relevant outcomes including patient-reported outcomes, physical functioning, and objective PA (via the Garmin Vivosmart4) were all possible during this study, with completion rates above the pre-specified 70%. Participants indicated that their interest in seeing results was a key motivator for completing the assessments. In terms of accuracy of the assessments, when available, it may be beneficial to perform physical functioning assessments in-person, as done in previous synchronous online delivered interventions^16^. The reliability and validity of the tests of physical function performed via videoconferencing have yet to be assessed, and some participants in the current study mentioned concerns about the validity of some of the measures used. Given the restrictions enforced by COVID-19, in-person assessments were not possible in the current study.

The objective measurement of PA was of particular interest during this pilot RCT. Commercially available wrist-worn activity trackers are becoming increasingly popular among the public^35^, and we found that data collection using these devices over a timeframe of up to 12 weeks was feasible. During this study, we developed protocols to enhance data collection. Firstly, additional time and information for participants were needed before study commencement to set up the activity tracker to increase wear time in the first week. Secondly, the trend of recording lower wear time in the final week, which has been reported previously in exercise oncology^21^, could be improved by sending reminders to synchronize the device before returning it to the study team. An additional consideration for using commercially available wearables devices in research is the valid interpretation of the data. Commercial companies usually have proprietary algorithms for their activity trackers (e,g, for MVPA minutes), and it can be difficult for researchers to access the raw dataset^36^. Researchers that are making interpretations based on data from commercially available trackers are therefore urged to make their analyses or algorithms publicly available to be refined and used in future trials. Bearing in mind the potential for commercially available trackers in healthcare, refining and sharing analyses to interpret the data from these devices will lead to wider use and therefore stronger objective evidence in exercise oncology.

Given the pilot feasibility nature of this work, limitations include recruiting predominately urban living participants, whereas a fully powered RCT will more likely recruit rural living participants. In addition, the intervention was administered early in the COVID-19 pandemic, and recruitment and adherence to online programs may differ once in-person options resume. Other limitations that should be accounted for in a future fully powered trial is the homogeneity of the sample (mainly woman diagnosed with breast cancer), the small sample size (meaning it is not possible to draw conclusions on our exploratory outcomes, see Supplementary File 4), and the fact that participants had completed an earlier online or in-person exercise oncology program (i.e., baseline ACE program). Addressing these in a future trial would increase the generalizability of a fully powered RCT’s findings to the larger population of people living with and beyond cancer. Given the potential of HC to support exercise interventions and the importance of targeting PA maintenance to enhance well-being in individuals living with and beyond cancer, further research on testing the effectiveness of such programming is needed^37,38^. Overall, while a need for online programming has been heightened during the COVID-19 pandemic ^4^, access to exercise oncology resources that support wellness for rural and remote populations, in particular, remains critical to address (EXCEL: NCT04478851).

### Conclusion

Synchronous online delivery of a supervised group-based exercise oncology maintenance program with additional HC support is feasible. The effectiveness of this intervention to aid individuals living with and beyond cancer in maintaining an active lifestyle and thereby improving physical functioning and QoL needs further investigation. Based on our findings, a fully powered trial is feasible and should offer at least two structured classes per week, give clear instructions on the use of an objective activity tracker, adapt questionnaires to the participants’ situation where possible, and provide meaningful results to the participants about their physical functioning.

## Supporting information

S1: The ACE Online Maintenance Program

S2: Health Coaching Protocol

S3: Exploratory Outcome Measures and Analysis

S4: Exploratory Results

S5: CONSORT 2010 Checklist

## Data Availability

All data produced in the present study are available upon reasonable request to the authors

## Authors’ contributions

**Max Eisele:** Conceptualization, Methodology, Software, Formal analysis, Investigation, Data curation Writing - Original Draft, Visualization, Project Administration **Rosie Twomey:** Methodology, Writing - Review & Editing, Supervision **Andrew J. Pohl:** Software, Resources, Data curation, Writing - Review & Editing **Meghan H. McDonough:** Methodology, Supervision, Writing - Review & Editing **Margaret S. McNeely:** Data curation, Writing - Review & Editing **Manuel Ester:** Investigation, Validation, Writing - Review & Editing **Julia Daun:** Investigation, Writing - Review & Editing **Nicole Culos-Reed:** Conceptualization, Methodology, Validation, Resources, Data curation, Writing - Review & Editing, Supervision, Funding acquisition

## Acknowledgments

We would like to thank the ACE participants for taking part in this study and acknowledge the contributions to this work by Tanya Williamson, Nicole Struthers, Emma McLaughlin, and Kelsey Ellis.

## Data and supplementary materials

S1: The ACE Online Maintenance Program

S2: Health Coaching Protocol

S3: Exploratory Outcome Measures and Analysis

S4: Exploratory Results

S5: CONSORT 2010 Checklist

## Footnotes

* Offered only in the 12-week HC protocol

