## Supplementary material for "The online delivery of exercise oncology classes supported with health coaching: A pilot randomized controlled trial": S1: The ACE Online Maintenance Program

Supplementary File 1

Online Exercise Class Intervention:

The classes were instructed by a CEP through Zoom video conferencing and were focused on strength and cardiovascular fitness The exercise classes consisted of 3-4 exercise circuits (i.e., series of exercises), with the last circuit focused on core stabilization. Each circuit was repeated twice and consisted of an exercise focused on a major muscle group, an exercise focused on a minor muscle group, and a cardiovascular focused activity. The exercise difficulty and amount were increased over the course of the intervention. Each exercise circuit was designed so that no exercise equipment was required. Alternative options and modifications to exercises were provided by the instructor, and exercises were tailored based on the individual's exercise experience, movement limitations, and equipment availability. Each class was monitored by a moderator (trained in exercise oncology), assisting the instructor by watching for participant safety. To ensure safety in the home setting and the ability to respond immediately to emergencies, participants were instructed to inform the moderator when they were moving off-camera, and participants provided their address and emergency contact information at study start. Before each exercise class, a reminder email with the upcoming class circuits and the Zoom link was sent out to the participants. Each class was an hour long, with 10 minutes of warmup, 40 minutes of exercise circuits, and 10 minutes of stretching at the end. Fifteen minutes before each class, the instructor was available for exercise-specific questions. After each class, the moderator facilitated a post-class discussion intended to foster social support, evoke thoughts about an active lifestyle, and offer the opportunity for questions. The first author and the exercise instructors met each week to discuss the fidelity of the protocol, potential considerations for individual participants, and discuss the upcoming exercise class plan. The classes were offered twice a week for the first two weeks of the program and then reduced to once a week for the remainder of the study. The tapered design of the exercise classes was chosen to first build familiarity with the exercises in a home setting, and then foster the maintenance of unsupervised PA through self-motivation to complete additional independent exercise prescription. No restrictions to engaging in other structured or unstructured physical activity classes were made.

*Unsupervised Home-based Program*

To support the participants in exercising on their own, a home-based exercise program prescription with embedded video links was sent to all participants at the beginning of Week 3. This program consisted of six different circuits with three different intensity levels (two low intensity; two moderate intensity; two high intensity) based on the traffic light system. The program was designed to promote self-efficacy, giving the participant the opportunity to control the intensity, duration and type of exercise based on their needs and daily fluctuations in energy and fatigue.

*Activity Tracker*

All participants received a commercially available activity tracker (Garmin Vivosmart4; Garmin Ltd. Kansas City, KS). Participants were instructed to download the Garmin Connect application (Garmin Ltd. Kansas City, KS) and log in with a study account to connect the account with the tracker. Participants were instructed to wear the tracker during waking hours and were allowed to use the Garmin to whatever extent they desired. The Garmin tracker was used to collect an objective measure of PA, and device usage was assessed.

Table 1. Behaviour change techniques (BCT) incorporated in the online maintenance program

| BCT domain | BCTs |
| --- | --- |
| 2. Feedback and monitoring |  |
|  | 2.1 Monitoring of behaviour by others without feedback |
|  | 2.3 Self-Monitoring of behaviour |
| 3. Social Support |  |
|  | 3.1 Social support (unspecified) |
|  | 3.2 Social support (practical) |
|  | 3.3 Social support (emotional) |
| 4. Shaping knowledge |  |
|  | 4.1 Instruction on how to perform a behaviour |
| 5. Natural consequences |  |
|  | 5.4 Monitoring of emotional consequences |
| 6. Comparison of behaviour |  |
|  | 6.1 Demonstration of the behaviour |
| 8. Repetition and substitution |  |
|  | 8.7 Graded tasks |
| 9. Comparison of outcomes |  |
|  | 9.1 Credible source |
| 10. Reward and threat |  |
|  | 10.4 Social reward |
| 12. Antecedents |  |
|  | 12.5 Adding objects to the environment |

*BCTs are based on Michie et al. (2013) Behaviour Change Taxonomy

Michie, S., Richardson, M., Johnston, M., Abraham, C., Francis, J., Hardeman, W., Eccles, M. P., Cane, J., & Wood, C. E. (2013). The behavior change technique taxonomy (v1) of 93 hierarchically clustered techniques: building an international consensus for the reporting of behavior change interventions. *Annals of Behavioral Medicine*, *46*(1), 81-95.
