## Supplementary material for "The online delivery of exercise oncology classes supported with health coaching: A pilot randomized controlled trial": S2: Health Coaching Protocol

Supplementary File 2

Health Coaching:

Participants randomized to the HC intervention received weekly individual HC calls. HC was structured based on Wolever et al.’s (2013) definition, which emphasizes that HC has to be participant-centred, built on a coach-participant relationship, and include participant-determined goals, a self-discovery process to find solutions, patient accountability, and education. A day before each HC call, the participants received a short survey on fatigue, QoL, stress, loneliness, and social support, enabling tailoring of the HC call to the individual.

Educational topics within the HC calls included: Goal Setting, Monitoring Behaviour, Barrier Management, Social Support, Stress Management, Adapting the Program, Self-compassion, Sleep & Nutrition, Reflection, Health Media, Remote Resources, and Maintaining Motivation. The order of the educational topics could be adjusted based on the specific participants’ needs each week. Based on the interests of the HC calls in the 8-week wave, the educational topics Self-Compassion, Sleep & Nutrition, Reflection, and Health Media were added to the 12-week wave. Each HC call was structured, starting off with a reflection on the previous week, a conversation about the educational topic, and finishing with an action plan for the upcoming week. Based on participant interest, a summary sheet of the educational topic was sent to the individual. At the half-way point of the intervention, the participant provided feedback on the HC calls, ensuring optimization of HC. The health coaches were graduate students trained in behaviour change strategies, exercise oncology, had extensive experience with the larger ACE program, and completed at least 30 hours of HC specific training (including mock interviews, motivational interviewing materials, and a literature review). The weekly HC calls were held via Zoom and at a convenient time for the participant. The length of each call was dependent on the participant’s needs. No restrictions on receiving additional counselling or coaching from outside sources were made.

Table 1. Behaviour change techniques (BCT) incorporated in the online maintenance program

| BCT domain | BCTs |
| --- | --- |
| 1. Goal setting and planning |  |
|  | 1.1 Goal setting (behaviour) |
|  | 1.2 Problem solving |
|  | 1.3 Goal setting (outcome) |
|  | 1.4 Action planning |
|  | 1.5 Review behaviour goals |
|  | 1.6 Discrepancy between current behaviour and goal |
|  | 1.7 Review outcome goals |
|  | 1.9 Commitment |
| 2. Feedback and monitoring |  |
|  | 2.2 Feedback on behaviour |
|  | 2.3 Self-Monitoring of behaviour |
|  | 2.4 Self-monitoring of outcome of behaviour |
|  | 2.7 Feedback on outcome of behaviour |
| 3. Social Support |  |
|  | 3.1 Social support (unspecified) |
|  | 3.2 Social support (practical) |
|  | 3.3 Social support (emotional) |
| 4. Shaping knowledge |  |
|  | 4.1 Instruction on how to perform a behaviour |
| 5. Natural consequences |  |
|  | 5.1 Information about health consequences |
|  | 5.4 Monitoring of emotional consequences |
|  | 5.6 Information about emotional consequences |
| 6. Comparison of behaviour |  |
|  | 6.1 Demonstration of the behaviour |
| 8. Repetition and substitution |  |
|  | 8.2 Behavior substitution |
|  | 8.6 Generalization of a target behaviour |
|  | 8.7 Graded tasks |
| 9. Comparison of outcomes |  |
|  | 9.1 Credible source |
| 10. Reward and threat |  |
|  | 10.4 Social reward |
| 13. Identity |  |
|  | 13.2 Framing/ reframing |
| 15. Self-belief |  |
|  | 15.1 Verbal persuasion about capability |
|  | 15.3 Focus on past successes |

*BCTs are based on Michie et al. (2013) Behaviour Change Taxonomy
