## Supplementary material for "The online delivery of exercise oncology classes supported with health coaching: A pilot randomized controlled trial": S3: Exploratory Outcome Measures and Analysis

Supplementary File 3

**EXPLORATORY OUTCOME MEASURES**

**Moderate to Vigorous Physical Activity Minutes**

MVPA was assessed objectively and through self-report. The modified Godin Leisure Time Exercise Questionnaire (mGLTEQ) ^1^ was used to assess self-reported MVPA per week at pre and post intervention. The Garmin Vivosmart4 was used as an objective measure of weekly MVPA throughout the study based on Garmin’s proprietary algorithm. To be considered for weekly objective MVPA calculation, the activity tracker had to have been worn for at least four valid days. A valid day was judged by wearing the tracker for at least 10h a day ^2,3^ with non-wear time being defined as 60 consecutive minutes ^4^. If the tracker was worn more than four days but not the complete seven days, the days of non-wear time were imputed by taking the average of the valid recorded days within that week. Adherence to exercise guidelines was determined by comparing the weekly objective minutes of MVPA to the oncology exercise guidelines of 90-minutes of MVPA per week^5^.

**Physical functioning**

Physical functioning assessments were administered via Zoom before (week 0) and after the intervention (week 9 or week 13 in the two waves, respectively) by a CEP blind to intervention allocation. The ACE testing protocol (McNeely et al., 2020) was modified to ensure safety during the online assessment and account for limitations in equipment in a home setting. Specifically, the cardiovascular fitness assessment was a 2-minute step test (instead of the 6-minute walk test used in ACE), the lower extremity flexibility assessment was a chair sit and reach test (instead of the box sit and reach test), and height and weight were self-reported (instead of measured by the research staff). Tests removed from the in-person protocol included grip strength and waist and hip circumference. Measures that followed the same procedures as during the in-person assessment included upper extremity flexibility (shoulder range of motion), balance (a one-legged stance with open eyes), and muscular endurance (30 second sit-to-stand; McNeely et al., 2020).

Upper extremity flexibility

Shoulder range of motion (ROM) was assessed by asking the participant to position themselves parallel to the device being used for the Zoom call, lift the arm closest to the screen up in a straight line parallel to the sagittal plane, and stop at the furthest point without arching their back. A screenshot was taken at the furthest ROM and analyzed after the assessment with a goniometer. Each shoulder was measured twice, and the average score of each side was recorded.

Lower extremity flexibility

Hamstring flexibility was assessed through a seated one-leg extension and reach test with a yardstick or tape measure. The participant was given instructions to hold a hamstring stretch for 30 seconds on each side to familiarize them with the test and reduce the risk of injury. For the test, the protocol outlined by Jones, Rikli, Max and Noffal ^6^ was followed. The only deviation from Jones et al.’s protocol was that the participant or a family member measured the distance to or from the toes while being instructed and observed by the assessor. The participant had two attempts, and the highest attempt was recorded to the nearest 0.5 cm. The criterion validity of this test compared to the reference test (goniometer measurement of a passive straight leg raise) was 0.76 and 0.81 for males and females, respectively. The intraclass test-retest reliability was excellent, with 0.92 and 0.96 for males and females, respectively ^6^.

Balance

Balance was assessed through an eyes-open one-legged stand with a cap at 45 seconds that is validated and reliable ^7^. For the test the protocol outlined by Franchignoni et al. (1998) was followed. Each leg was assessed once, and the time was recorded to the nearest 0.1 second ^8^.

Muscular Endurance

Muscular endurance was measured through the validated, and reliable 30-seconds sit-to-stand test ^9,10^. The test measures the times a subject can get up from a seated chair position in 30 seconds. The protocol outlined by Jones et al. was followed ^9^.

Cardiovascular Fitness

Cardiovascular fitness was assessed through the 2-minute step test. In this test the number of steps with the knee above the midway point from the patella to iliac crest in 2 minutes was counted. The protocol outlined by Rikli and Jones was followed ^11^. The participant was asked to complete the test perpendicular to the camera and ideally next to a background that allowed for easy detection of each leg during the Zoom call. This measure is commonly used in older adults and has a moderate validity to assess cardiovascular fitness ^12^.

**Patient Reported Outcomes**

Patient-reported outcomes were administered at baseline and post-intervention through a closed survey on SurveyMonkey. Participants received an email with a unique survey link. The survey consisted of 78 questions, was optional, and did not include any incentives for completion. Participants’ responses were de-identified by replacing their registration email addresses with study IDs for analysis. The description of the surveys used was based on the Checklist for Reporting Results of Internet E-Surveys (CHERRIES) ^13^.

Quality of Life (QOL)

QOL was assessed through the functional assessment of cancer therapy general scale (FACT-G). The change in the overall FACT-G score as well as the score of each domain subscale (physical well-being, social/family well-being, emotional well-being, and functional well-being) was evaluated. This tool has shown to be simple and fast to complete while still having high reliability and validity across cancer age groups ^14,15^.

Fatigue

Fatigue was assessed through the functional assessment of chronic illness therapy fatigue scale (FACIT-F). The 13-item fatigue subscale has been commonly used in the cancer population and is reliable and valid ^16^.

Barrier Self-Efficacy

Barrier self-efficacy was assessed using a scale developed based on the most common barriers for cancer survivors ^17^. The scale consists of nine barriers, and the participant was asked how confident they are to exercise despite these barriers. The scale has a high internal consistency and good test-retest reliability ^17^.

COVID-19 Impact

Stress, loneliness, and social support were assessed through the Perceived Stress Scale (PSS), the short form of the UCLA Loneliness Scale (ULS-6), and the Oslo Social Support Scale (OSSS-3), respectively. The 10-item PSS is a simple and widely established scale that has acceptable psychometric properties ^18^. The 6-item ULS-6 is a brief, reliable, and valid measure of loneliness ^19^. The 3-item OSSS-3 is a short, reliable and valid measure of social support ^20^.

**EXPLORATORY ANALYSIS**

All data were analyzed using SPSS statistics (v26, IBM). For continuous data (age, BMI, MVPA, TMST, and change scores), normality was assessed by inspecting histograms, box plots, QQ-plots, the Shapiro-Wilk test of normality. Descriptive statistics are reported as mean and standard deviation (SD) for normally distributed data and medians and interquartile ranges (IQR) for non-normal distributed data. Categorical outcomes are reported as frequency and percentage. For pre-post intervention data, mean change scores were presented with 95% confidence intervals (95% CI) ^44^. Where available, change scores were compared with minimal clinically importance differences (MCIDs); the smallest difference that is perceived by patients as beneficial or harmful ^45^. The proportion of participants improving or deteriorating by more than the MCID is also reported. Effect sizes (standardized mean difference) were calculated (Cohen’s *d* or Hedge’s *g*) and interpreted as 0.2 is small, 0.5 is moderate and 0.8 is large, in line with Cohen (1992).
