## Supplementary material for "The online delivery of exercise oncology classes supported with health coaching: A pilot randomized controlled trial": S4: Exploratory Results

Supplementary File 3

Percentage Weartime

| Intervention Week | Wave 1 | Wave 2 |
| --- | --- | --- |
| 1 |  | 78.022% |
| 2 |  | 89.7959% |
| 3 |  | 85.7143% |
| 4 |  | 86.7347% |
| 5 | 78.882% | 85.7143% |
| 6 | 81.9876% | 87.7551% |
| 7 | 80.1242% | 85.7143% |
| 8 | 68.9441% | 86.7347% |
| 9 |  | 79.5918% |
| 10 |  | 82.6531% |
| 11 |  | 90.8163% |
| 12 |  | 76.5306% |

Figure 1. Percent Wear time per Week in the 8-Week and the 12-Week Wave

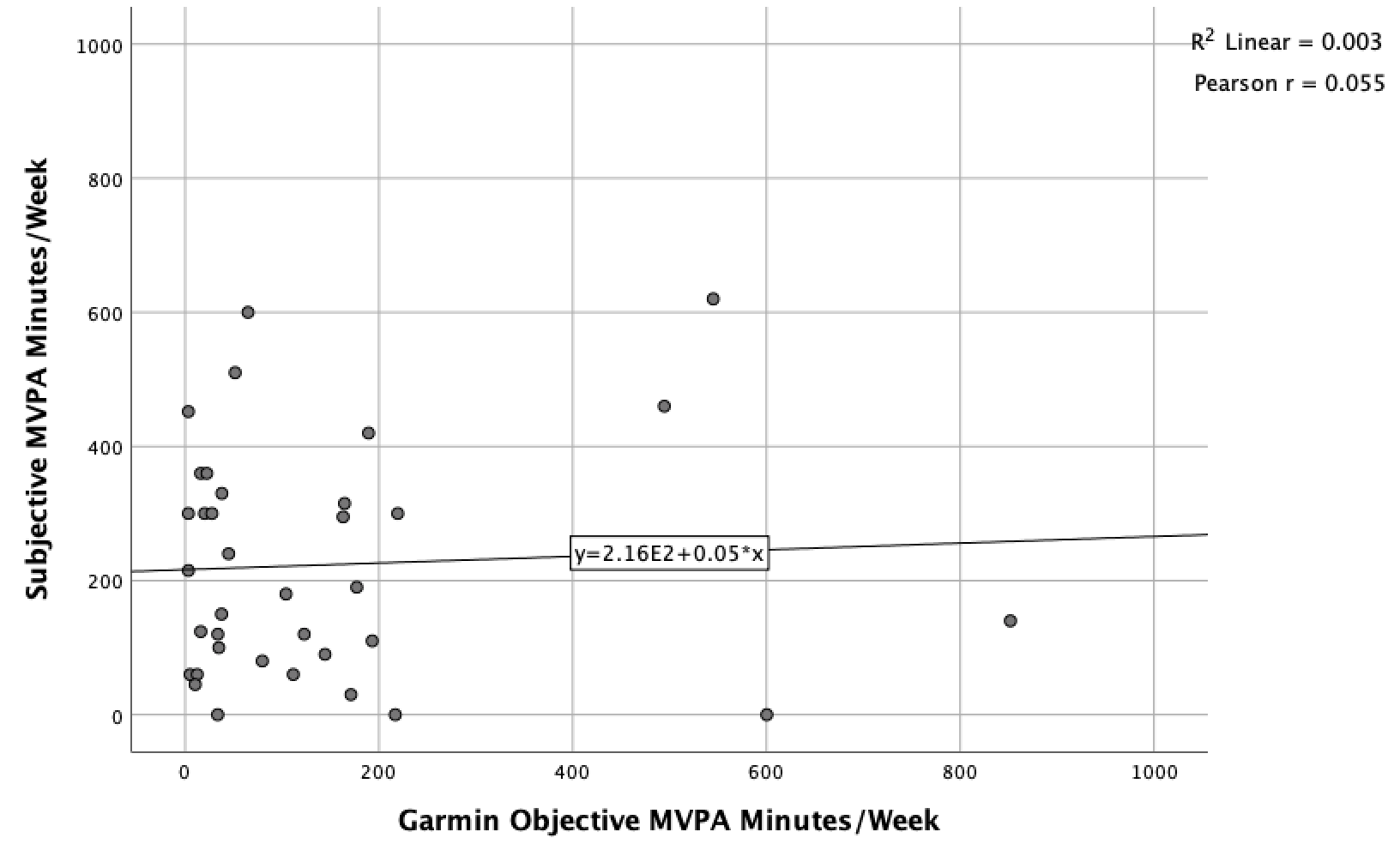

Figure 2. Correlation between the Subjective modified Godin Leisure Time Exercise Questionnaire (mGLTEQ) and the Objective Garmin Vivosmart4 to measure Moderate to Vigorous Physical Activity (MVPA) Minutes per Week

**Table 1.** **Baseline to Post-intervention Differences for the non-HC Intervention Group in the 12-week Wave**

|  | Baseline:  Mean ± SD  Median (IQR) | 12WK  Mean ± SD  Median (IQR) | Mean ± SD /  Median (IQR) Difference | 95% CI | ES:  *d* |
| --- | --- | --- | --- | --- | --- |
| FACT-G | 71 ± 9 | 79 ± 8 | 8 ± 8 | 2.68 – 13.32 | 0.93 |
| FACIT-F | 36 ± 10 | 39 ± 6 | 3 ± 8 | -3.20 – 8.70 | 0.33 |
| Barrier SE | 45 (37) | 45 (22) | -1 ± 20 | -21.55 – 21.79 | 0.00 |
| OSSS-3 | 11 ± 2 | 11 ± 3 | 1 ± 2 | -0.69 – 2.19 | 0.33 |
| PSS | 19 ± 4 | 15 ± 6 | -4 ± 7 | 0.76 – 7.48 | 0.82 |
| ULS-6 | 15 (5) | 14 (8) | -2 (5) | -3.86 – 5.86 | 0.15 |
| MVPA (min/week) | 250.6 ± 166.1 | 219.4 ± 186.8 | -31.3 ± 180.8 | -79.23 – 141.73 | 0.18 |
| BMI kg/m^2^ | 26.6 (7.7) | 26.5 (7.5) | -0.3 ± 1.1 | -4.58 – 4.84 | 0.02 |
| SROM (R.) (degree) | 149.5 ± 12.0 | 151.0 ± 14.0 | 3.0 ± 14.5 | -6.61 – 10.01 | 0.13 |
| SROM (L.) (degree) | 157.0 (7.0) | 157.5 (7.5) | 4.0 (6.5) | -3.96 – 4.96 | 0.07 |
| Steps (reps) | 115 ± 23 | 116 ± 20 | -2.5 (28.5) | -11.91 – 14.91 | 0.07 |
| Sit-to-Stand (reps) | 15 ± 2 | 15 ± 2 | 0 ± 1.5 | -1.25 – 1.85 | 0.13 |
| Reach R. (cm) | 2.5 (10.5) | 9.5 (19.0) | 3.5 (7.5) | -4.15 – 18.35 | 0.46 |
| Reach L. (cm) | 2.5 (13.0) | 8.5 (21.0) | 2.5 (6.5) | -6.31 – 18.11 | 0.34 |
| Balance R (sec) | 36.5 (30.2) | 39.1 (27.4) | 0.00 (4.7) | -10.57 – 25.37 | 0.26 |
| Balance L (sec) | 41.1 (28.3) | 45.0 (28.1) | 0.00 (18.5) | -13.57 – 21.37 | 0.14 |

ES=Effect size; Cohen’s d of 0.2, 0.5, and 0.8 was judged as a small, moderate, and large effect size (Cohen, 1992)

FACT-G = Functional Assessment of Cancer Therapy – General (QoL); FACIT-F = Functional Assessment of Chronic Illness Therapy – Fatigue; Barrier SE = Barrier Self-Efficacy; OSSS-3 = Oslo Social Support Scale; PSS = Perceived Stress Scale; ULS-6 = UCLA Loneliness Scale; MVPA = Moderate to Vigorous Physical Activity; BMI = Body Mass Index; SROM = Shoulder Range of Motion; Reach = Sit and Reach test

**Table 2.** **Baseline to Post-intervention Differences for the HC Intervention Group in the 12-week Wave**

|  | Baseline:  Mean ± SD  Median (IQR) | 12WK  Mean ± SD  Median (IQR) | Mean ± SD /  Median (IQR) Difference | 95% CI | ES:  d  /  (g) |
| --- | --- | --- | --- | --- | --- |
| FACT-G | 81 ± 7 | 81 ± 8 | 0 ± 6 | -4.66 – 5.52 | 0.06 |
| FACIT-F | 41 ± 4 | 41 ± 4 | 0 ± 4 | -2.77 – 3.05 | 0.03 |
| BarrierSE | 53 (23) | 66 (33) | 4 ± 28 | -8.02 – 34.02 | 0.46 |
| OSSS-3 | 11 ± 1 | 11 ± 2 | 0 ± 2 | -0.95 – 1.23 | 0.09 |
| PSS | 13 ± 6 | 15 ± 4 | 2 ± 5 | -1.71 – 5.71 | 0.40 |
| ULS-6 | 15 (6) | 16 (7) | -1 (1) | -3.54 – 5.54 | 0.15 |
| MVPA (min/week) | 165.0 ± 132.9 | 229.1 ± 150.8 | 64.1 ± 101 | -34.27 – 162.55 | 0.45 |
| BMI kg/m^2^ | 24.5 (4.1) | 25.0 (5.1) | -0.1 ± 0.6 | -2.79 – 3.73 | 0.10 |
| SROM (R.) (degree) | 161.0 ± 10 | 157.5 ± 6.5 | -1.5 ± 7.0 | -3.16 – 9.76 | 0.39 |
| SROM (L.) (degree) | 151.0 (6.0) | 162.5 (7.0) | -0.5 (18.5) | 6.96 – 16.04 | 1.76 |
| Steps (reps) | 111 ± 9 | 119 ± 12 | 9 (9) | 0.01 – 15.79 | 0.72 |
| Sit-to-Stand (reps) | 23 ± 7 | 24 ± 7 | 1 ± 5 | -4.72 – 5.72 | (0.07) |
| Reach R. (cm) | 12.0 (8.5) | 15.5 (13.0) | 3.5 (5.0) | -4.84 – 11.84 | 0.32 |
| Reach L. (cm) | 9.5 (5.5) | 14.5 (13.0) | 3.0 (5.5) | -4.04 – 14.04 | 0.50 |
| Balance R (sec) | 45.0 (8.7) | 45.0 (0.0) | 0.0 (8.7) | -8.05 – 8.05 | 0.00 |
| Balance L (sec) | 45.0 (0.0) | 45.0 (0.0) | 0.0 (0.0) | 0.00 – 0.00 | 0.00 |

ES=Effect size; Cohen’s d of 0.2, 0.5, and 0.8 was judged as a small, moderate, and large effect size (Cohen, 1992)

FACT-G = Functional Assessment of Cancer Therapy – General (QoL); FACIT-F = Functional Assessment of Chronic Illness Therapy – Fatigue; Barrier SE = Barrier Self-Efficacy; OSSS-3 = Oslo Social Support Scale; PSS = Perceived Stress Scale; ULS-6 = UCLA Loneliness Scale; MVPA = Moderate to Vigorous Physical Activity; BMI = Body Mass Index; SROM = Shoulder Range of Motion; Reach = Sit and Reach test

**Table 3. Baseline to Post-intervention Differences for the 12-Week Wave**

| Outcome Measure | Baseline  (Mean ± SD  Median (IQR)) | 12WK  (Mean ± SD  Median (IQR)) | 95% CI | ES:  d  or  (g) |
| --- | --- | --- | --- | --- |
| FACT-G | 76 ± 9 | 80 ± 8 | 0.86 – 8.06 | 0.52 |
| FACIT-F | 40 (9) | 40 (7) | -3.44 – 3.44 | 0.00 |
| Barrier SE | 52 ± 21 | 53 ± 19 | -6.95 – 9.89 | 0.10 |
| OSSS-3 | 11 (2) | 11 (1) | -4.25 – 4.25 | 0.00 |
| PSS | 16 ± 6 | 15 ± 5 | -0.92 – 3.44 | 0.24 |
| ULS-6 | 15 ± 3 | 14 ± 4 | -0.61 – 2.21 | 0.25 |
| MVPA (min/week) | 210.7 ± 152.7 | 223.9 ± 165.0 | -52.22 – 78.74 | 0.08 |
| BMI kg/m^2^ | 26.7 ± 5.3 | 26.5 ± 5.2 | -1.95 – 2.37 | 0.04 |
| SROM (R.) (degree) | 155.0 ± 12.5 | 154.0 ± 11.5 | -4.21 – 5.55 | 0.06 |
| SROM (L.) (degree) | 156.0 (10.0) | 158.5 (10.5) | -1.71 – 6.71 | 0.24 |
| Steps (reps) | 113 ± 17 | 118 ± 17 | -2.47 – 11.39 | 0.26 |
| Sit-to-Stand (reps) | 16 (9) | 18 (10) | -2.60 – 5.60 | (0.16) |
| Reach R. (cm) | 5.0 (12.0) | 12.5 (15.0) | 1.75 – 13.25 | 0.55 |
| Reach L. (cm) | 8.0 (12.0) | 12.0 (19.5) | -3.53 – 13.59 | 0.25 |
| Balance R (sec) | 45.0 (17.1) | 45.0 (7.7) | -7.02 – 7.02 | 0.00 |
| Balance L (sec) | 45.0 (12.1) | 45.0 (0.0) | -6.70 – 6.70 | 0.00 |

ES=Effect size; Cohen’s d / Hedge’s g of 0.2, 0.5, and 0.8 was judged as a small, moderate, and large effect size (Cohen, 1992)

FACT-G = Functional Assessment of Cancer Therapy – General (QoL); FACIT-F = Functional Assessment of Chronic Illness Therapy – Fatigue; Barrier SE = Barrier Self-Efficacy; OSSS-3 = Oslo Social Support Scale; PSS = Perceived Stress Scale; ULS-6 = UCLA Loneliness Scale; MVPA = Moderate to Vigorous Physical Activity; BMI = Body Mass Index; SROM = Shoulder Range of Motion; Reach = Sit and Reach test

**Table 4. Change Scores of the HC and Non-HC group for the 12-Week Wave**

| Outcome Measure | Non-HC:  Mean ± SD  Median (IQR) | HC:  Mean ± SD  Median (IQR) | 95% CI | ES:  g |
| --- | --- | --- | --- | --- |
| FACT-G | 8 ± 8 | 0 ± 6 | 1.51 - 13.63 | 1.04 |
| FACIT-F | 3 ± 8 | 0 ± 4 | -4.53 – 10.31 | 0.44 |
| Barrier SE | -1 ± 20 | 4 ± 28 | -24.80 – 15.30 | 0.20 |
| OSSS-3 | 1 ± 2 | 0 ± 2 | -0.74 – 1.96 | 0.37 |
| PSS | -4 ± 7 | 2 ± 5 | -10.85 – -1.41 | 1.08 |
| ULS-6 | -2 (5) | -1 (1) | -4.87 – 3.87 | 0.13 |
| MVPA (min/week) | -31.3 ± 180.8 | 64.1 ± 101 | -218.03 – 27.25 | 0.64 |
| BMI kg/m^2^ | -0.3 ± 1.1 | -0.1 ± 0.6 | -1.04 – 0.46 | 0.32 |
| SROM (R.) (degree) | 3.0 ± 14.5 | -1.5 ± 7.0 | -5.17 – 13.67 | 0.37 |
| SROM (L.) (degree) | 4.0 (6.5) | -0.5 (18.5) | -10.53 – 19.53 | 0.33 |
| Steps (reps) | -3 (28) | 9 (9) | -35.67 – 12.67 | 0.53 |
| Sit-to-Stand (reps) | 0 ± 2 | 1 ± 5 | -3.53 – 4.53 | 0.15 |
| Reach R. (cm) | 3.5 (7.5) | 3.5 (5.0) | -7.23 -7.23 | 0.00 |
| Reach L. (cm) | 2.5 (6.5) | 3.0 (5.5) | -7.61 – 6.11 | 0.12 |
| Balance R (sec) | 0.0 (4.7) | 0.0 (8.7) | -7.64 – 7.64 | 0.00 |
| Balance L (sec) | 0.0 (18.5) | 0.0 (0.0) | -15.15 – 15.15 | 0.00 |

ES=Effect size; Cohen’s d / Hedge’s g of 0.2, 0.5, and 0.8 was judged as a small, moderate, and large effect size (Cohen, 1992)

FACT-G = Functional Assessment of Cancer Therapy – General (QoL); FACIT-F = Functional Assessment of Chronic Illness Therapy – Fatigue; Barrier SE = Barrier Self-Efficacy; OSSS-3 = Oslo Social Support Scale; PSS = Perceived Stress Scale; ULS-6 = UCLA Loneliness Scale; MVPA = Moderate to Vigorous Physical Activity; BMI = Body Mass Index; SROM = Shoulder Range of Motion; Reach = Sit and Reach test

Table 5. Percent of Participants Improving, Maintaining, or Worsening by the Minimal Clinically Important Difference for the 12-Week Wave

|  | Improved | | Maintained | | Worsen | |
| --- | --- | --- | --- | --- | --- | --- |
|  | HC  *n* (%) | Non-HC  *n* (%) | HC  *n* (%) | Non-HC  *n* (%) | HC  *n* (%) | Non-HC  *n* (%) |
| FACT-G | 2 (28.6) | 6 (75.0) | 3 (42.9) | 1 (12.5) | 2 (28.6) | 1 (12.5) |
| FACIT-F | 2 (28.6) | 3 (37.5) | 4 (57.1) | 3 (37.5) | 1 (14.3) | 2 (25.0) |
| Balance(R) | 0 (0) | 0 (0) | 7 (100.0) | 8 (100.0) | 0 (0) | 0 (0) |
| Balance(L) | 0 (0) | 1 (12.5) | 7 (100.0) | 7 (87.5) | 0 (0) | 0 (0) |
| SROM (R) | 0 (0) | 0 (0) | 7 (100.0) | 8 (100.0) | 0 (0) | 0 (0) |
| SROM (L) | 1 (14.3) | 1 (12.5) | 6 (85.7) | 7 (87.5) | 0 (0) | 0 (0) |
| MVPA | 5 (71.4) | 3 (37.5) | 0 (0) | 1 (12.5) | 2 (28.6) | 4 (50.0) |
| Sit-to-Stand | 1 (16.7) | 1 (12.5) | 4 (66.7) | 7 (87.5) | 1 (16.7) | 0 (0) |
| PSS | 2 (28.6) | 4 (50.0) | 2 (28.6) | 3 (37.5) | 3 (42.9) | 1 (12.5) |

MCIDs: FACT-G = 3 points; FACIT-F = 3 points (McNeely et al., 2020); Balance = 24 seconds (Goldberg et al., 2011); SROM = >10°; MVPA = 26min/week (Hur et al., 2019); Sit-to-Stand = 2.6 repetitions (Wright et al., 2011); PSS-10 = 2.66 points (Drachev et al., 2020)

FACT-G = Functional Assessment of Cancer Therapy – General (QoL); FACIT-F = Functional Assessment of Chronic Illness Therapy – Fatigue; PSS = Perceived Stress Scale; MVPA = Moderate to Vigorous Physical Activity; SROM = Shoulder Range of Motion;

*Missing Values: Sit-to-Stand = 1 (HC).

**Table 6.** **Baseline to Post-intervention Differences for the non-HC Intervention Group in the 8-Week Wave**

|  | Baseline:  Mean ± SD  Median (IQR) | 8WK  Mean ± SD  Median (IQR) | Mean ± SD /  Median (IQR) Difference | g | 95% CI |
| --- | --- | --- | --- | --- | --- |
| FACT-G | 78.3 ± 8.7 | 80.8 ± 7.3 | 1.17 ± 5.31 | 0.31 | -4.18 – 9.18 |
| FACIT-F | 36.8 ± 5.2 | 35.6 ± 12.1 | -1.08 ± 9.61 | 0.13 | -8.80 – 6.40 |
| Barrier SE | 46.00 (26.00) | 38.50 (35.00) | -5.50 (28.0) | 0.24 | -32.87 – 17.87 |
| OSSS-3 | 11.1 ± 2.4 | 10.8 ± 2.3 | -0.50 ± 1.62 | 0.13 | -2.25 – 1.65 |
| PSS | 15.0 (10.0) | 14.5 (11.5) | -1.08 ± 4.44 | 0.05 | -9.40 – 8.40 |
| ULS-6 | 13.2 ± 3.9 | 13.9 ± 2.8 | 1.08 ± 2.87 | 0.20 | -2.13 – 3.53 |
| MVPA (min/week) | 30.0 (225.0) | 120.0 (252.5) | 30.00 (120.00) | 0.38 | -108.54 – 288.54 |
| BMI kg/m^2^ | 27.6 ± 5.6 | 27.3 ± 6.5 | 0.11 (0.54) | 0.05 | -5.55 – 4.95 |
| SROM (R.) (degree) | 152.7 ± 8.3 | 152.1 ± 9.4 | -0.45 ± 4.12 | 0.07 | -8.09 – 6.89 |
| SROM (L.) (degree) | 151.2 ± 7.4 | 148.9 ± 6.7 | -1.82 ± 7.17 | 0.32 | -8.32 – 3.72 |
| Steps (reps) | 89.0 (23.3) | 98.0 (32.0) | 9.30 ± 10.23 | 0.32 | -15.12 – 33.12 |
| Sit to Stand (reps) | 17.7 ± 5.9 | 18.6 ± 7.4 | 0.40 ± 2.59 | 0.14 | -4.88 – 6.68 |
| Reach R. (cm) | 7.0 ± 12.1 | 7.9 ± 8.6 | 0.50 (7.50) | 0.08 | -8.15 – 9.95 |
| Reach L. (cm) | 6.7 ± 11.1 | 7.8 ± 8.4 | 1.50 (13.00) | 0.11 | -7.37 – 9.57 |
| Balance R (sec) | 38.9 (38.8) | 45.0 (30.6) | 0.00 (6.10) | 0.17 | -23.90 – 36.10 |
| Balance L (sec) | 45.0 (24.2) | 21.4 (34.8) | 0.00 (22.93) | 0.79 | -49.40 – 2.20 |

ES=Effect size; Hedge’s g of 0.2, 0.5, and 0.8 was judged as a small, moderate, and large effect size (Cohen, 1992)

FACT-G = Functional Assessment of Cancer Therapy – General (QoL); FACIT-F = Functional Assessment of Chronic Illness Therapy – Fatigue; Barrier SE = Barrier Self-Efficacy; OSSS-3 = Oslo Social Support Scale; PSS = Perceived Stress Scale; ULS-6 = UCLA Loneliness Scale; MVPA = Moderate to Vigorous Physical Activity; BMI = Body Mass Index; SROM = Shoulder Range of Motion; Reach = Sit and Reach test

**Table 7.** **Baseline to Post-intervention Differences for the HC Intervention Group in the 8-Week Wave**

|  | Baseline:  Mean ± SD  Median (IQR) | 8WK  Mean ± SD  Median (IQR) | Mean ± SD /  Median (IQR) Difference | ES:  d | 95% CI |
| --- | --- | --- | --- | --- | --- |
| FACT-G | 79.9 ± 14.1 | 84.8 ± 12.3 | 4.92 ± 6.87 | 0.37 | -1.40 – 11.20 |
| FACIT-F | 36.9 ± 9.5 | 40.9 ± 8.5 | 4.00 ± 6.93 | 0.44 | -0.28 – 8.28 |
| Barrier SE | 38.00 (43.00) | 61.00 (37.00) | 5.50 (42.75) | 0.57 | 3.84 – 42.16 |
| OSSS-3 | 11.0 ± 2.3 | 10.9 ± 2.1 | -0.08 ± 1.24 | 0.05 | -0.94 – 1.14 |
| PSS | 17.0 (12.8) | 13.5 (7.8) | -1.33 ± 4.74 | 0.33 | -2.18 – 9.18 |
| ULS-6 | 12.6 ± 4.0 | 13.3 ± 3.1 | 0.75 ± 1.71 | 0.20 | -1.05 – 2.45 |
| MVPA (min/week) | 90.0 (172.5) | 257.5 (309.0) | 85.00 (221.25) | 0.67 | 28.46- 306.54 |
| BMI kg/m^2^ | 27.4 ± 5.7 | 26.8 ± 5.4 | -0.33 (0.89) | 0.11 | -2.02 – 3.22 |
| SROM (R.) (degree) | 157.0 ± 13.4 | 157.0 ± 10.3 | 0.00 ± 6.64 | 0.00 | -5.87 – 5.87 |
| SROM (L.) (degree) | 158.6 ± 8.9 | 158.2 ± 7.5 | -0.42 ± 6.99 | 0.05 | -3.55 – 4.35 |
| Steps (reps) | 83.5 (16.5) | 96.5 (18.8) | 8.67 ± 12.24 | 0.73 | 4.58 – 21.42 |
| Sit to Stand (reps) | 16.8 ± 3.2 | 17.3 ± 4.8 | 0.42 ± 5.16 | 0.12 | -1.61 – 2.61 |
| Reach R. (cm) | 6.9 ± 8.7 | 9.3 ± 9.1 | 4.25 (5.38) | 0.27 | -1.80 – 6.60 |
| Reach L. (cm) | 6.8 ± 7.5 | 9.7 ± 7.4 | 4.00 (6.38) | 0.39 | -0.61 – 6.41 |
| Balance R (sec) | 45.0 (18.8) | 45.0 (16.5) | 0.00 (5.25) | 0.00 | -8.42 – 8.42 |
| Balance L (sec) | 45.0 (0.0) | 45.0 (25.0) | 0.00 (3.15) | 0.00 | -15.88 – 15.88 |

ES=Effect size; Cohen’s d of 0.2, 0.5, and 0.8 was judged as a small, moderate, and large effect size (Cohen, 1992)

FACT-G = Functional Assessment of Cancer Therapy – General (QoL); FACIT-F = Functional Assessment of Chronic Illness Therapy – Fatigue; Barrier SE = Barrier Self-Efficacy; OSSS-3 = Oslo Social Support Scale; PSS = Perceived Stress Scale; ULS-6 = UCLA Loneliness Scale; MVPA = Moderate to Vigorous Physical Activity; BMI = Body Mass Index; SROM = Shoulder Range of Motion; Reach = Sit and Reach test

**Table 8. Baseline to Post-intervention Differences for the 8-Week Wave**

| Outcome Measures | 8-Week Wave | | | |
| --- | --- | --- | --- | --- |
|  | Baseline  (Mean ± SD  Median (IQR)) | 8WK  (Mean ± SD  Median (IQR)) | ES:  g | 95% CI |
| FACT-G | 79.08 ± 11.87 | 82.79 ± 10.13 | 0.34 | -2.64 – 10.06 |
| FACIT-F | 36.84 ± 7.39 | 38.25 ± 10.61 | 0.15 | -3.83 – 6.65 |
| Barrier SE | 46.56 ± 23.06 | 48.88 ± 20.58 | 0.11 | -10.26 – 14.90 |
| OSSS-3 | 11.04 ± 2.32 | 10.83 ± 2.14 | 0.09 | -1.49 – 1.07 |
| PSS | 15.44 ± 6.61 | 14.00 ± 6.26 | 0.22 | -5.14 – 2.26 |
| ULS-6 | 12.88 ± 3.88 | 13.63 ± 2.89 | 0.22 | -1.22 – 2.72 |
| MVPA (min/week) | 90.00 (215.00) | 185.00 (234.00) | 0.42 | -34.07 – 224.07 |
| BMI kg/m^2^ | 27.50 ± 5.54 | 27.01 ± 5.76 | 0.09 | -3.81 – 2.83 |
| SROM (R.) (degree) | 154.74 ± 10.99 | 154.65 ± 9.97 | 0.01 | -6.21 – 6.03 |
| SROM (L.) (degree) | 154.74 ± 8.80 | 153.71 ± 8.41 | 0.12 | -6.04 – 3.98 |
| Steps (reps) | 88.46 ± 16.17 | 97.39 ± 19.42 | 0.50 | -1.55 – 19.41 |
| Sit to Stand (reps) | 17.25 ± 4.69 | 17.87 ± 6.10 | 0.11 | -2.57 – 3.81 |
| Reach R. (cm) | 6.94 ± 10.38 | 8.64 ± 8.67 | 0.18 | -3.88 – 7.28 |
| Reach L. (cm) | 6.76 ± 9.39 | 8.76 ± 7.79 | 0.23 | -3.04 – 7.04 |
| Balance R (sec) | 45.00 (30.00) | 45.00 (22.00) | 0.00 | -15.40 – 15.40 |
| Balance L (sec) | 45.00 (16.00) | 45.00 (32.8) | 0.00 | -15.27 – 15.27 |

ES=Effect size; Cohen’s d / Hedge’s g of 0.2, 0.5, and 0.8 was judged as a small, moderate, and large effect size (Cohen, 1992)

FACT-G = Functional Assessment of Cancer Therapy – General (QoL); FACIT-F = Functional Assessment of Chronic Illness Therapy – Fatigue; Barrier SE = Barrier Self-Efficacy; OSSS-3 = Oslo Social Support Scale; PSS = Perceived Stress Scale; ULS-6 = UCLA Loneliness Scale; MVPA = Moderate to Vigorous Physical Activity; BMI = Body Mass Index; SROM = Shoulder Range of Motion; Reach = Sit and Reach test

**Table 9. Change Scores of the HC and Non-HC group for both the 8-Week Wave**

| Outcome Measures | 8-Week Wave | | | |
| --- | --- | --- | --- | --- |
|  | Non-HC:  Mean ± SD  Median (IQR) | HC:  Mean ± SD  Median (IQR) | ES:  d  or  (g) | 95% CI |
| FACT-G | 1.17 ± 5.31 | 4.92 ± 6.87 | 0.61 | 0.74 – 6.76 |
| FACIT-F | -1.08 ± 9.61 | 4.00 ± 6.93 | 0.61 | 0.88 – 9.28 |
| Barrier SE | -5.50 (28.0) | 5.50 (42.75) | 0.30 | -7.79 – 29.79 |
| OSSS-3 | -0.50 ± 1.62 | -0.08 ± 1.24 | 0.29 | -0.29 – 1.13 |
| PSS | -1.08 ± 4.44 | -1.33 ± 4.74 | 0.05 | -1.92 – 2.42 |
| ULS-6 | 1.08 ± 2.87 | 0.75 ± 1.71 | 0.14 | -0.95 – 1.61 |
| MVPA (min/week) | 30.00 (120.00) | 85.00 (221.25) | 0.31 | -45.11 – 155.11 |
| BMI kg/m^2^ | 0.11 (0.54) | -0.33 (0.89) | (0.58) | -1.11 – 0.23 |
| SROM (R.) (degree) | -0.45 ± 4.12 | 0.00 ± 6.64 | (0.08) | -4.40 – 5.30 |
| SROM (L.) (degree) | -1.82 ± 7.17 | -0.42 ± 6.99 | (0.20) | -4.74 – 7.54 |
| Steps (reps) | 9.30 ± 10.23 | 8.67 ± 12.24 | (0.06) | -10.79 – 9.53 |
| Sit to Stand (reps) | 0.40 ± 2.59 | 0.42 ± 5.16 | (0.00) | -3.73 – 3.77 |
| Reach R. (cm) | 0.50 (7.50) | 4.25 (5.38) | (0.58) | -1.87 – 9.37 |
| Reach L. (cm) | 1.50 (13.00) | 4.00 (6.38) | (0.25) | -6.26 – 11.26 |
| Balance R (sec) | 0.00 (6.10) | 0.00 (5.25) | (0.00) | -4.92 – 4.92 |
| Balance L (sec) | 0.00 (22.93) | 0.00 (3.15) | (0.00) | -13.90 – 13.90 |

ES=Effect size; Cohen’s d / Hedge’s g of 0.2, 0.5, and 0.8 was judged as a small, moderate, and large effect size (Cohen, 1992)

FACT-G = Functional Assessment of Cancer Therapy – General (QoL); FACIT-F = Functional Assessment of Chronic Illness Therapy – Fatigue; Barrier SE = Barrier Self-Efficacy; OSSS-3 = Oslo Social Support Scale; PSS = Perceived Stress Scale; ULS-6 = UCLA Loneliness Scale; MVPA = Moderate to Vigorous Physical Activity; BMI = Body Mass Index; SROM = Shoulder Range of Motion; Reach = Sit and Reach test

Table 10. Percent of Participants Improving, Maintaining, or Worsening by the Minimal Clinically Important Difference for the 8-Week Wave

|  | Improved | | Maintained | | Worsen | |
| --- | --- | --- | --- | --- | --- | --- |
|  | HC  n (%) | Non-HC  n (%) | HC  n (%) | Non-HC  n (%) | HC  n (%) | Non-HC  n (%) |
| FACT-G | 6 (50.0) | 6 (50.0) | 4 (33.3) | 3 (25.0) | 2 (16.7) | 3 (25.0) |
| FACIT-F | 8 (66.7) | 6 (50.0) | 3 (25.0) | 1 (8.3) | 1 (8.3) | 5 (41.7) |
| Balance(R) | 1 (8.3) | 0 (0.0) | 11 (91.7) | 11 (100.0) | 0 (0.0) | 0 (0.0) |
| Balance(L) | 0 (0.0) | 0 (0.0) | 11 (91.7) | 9 (81.8) | 1 (8.3) | 1 (9.1) |
| SROM (R) | 0 (0.0) | 0 (0.0) | 12 (100.0) | 11 (100.0) | 0 (0.0) | 0 (0.0) |
| SROM (L) | 0 (0.0) | 0 (0.0) | 12 (100.0) | 11 (100.0) | 0 (0.0) | 0 (0.0) |
| MVPA | 9 (75.0) | 8 (66.7) | 2 (16.7) | 2 (16.7) | 1 (8.3) | 2 (16.7) |
| SittoStand | 3 (25.0) | 2 (20.0) | 7 (58.3) | 7 (70.0) | 2 (16.7) | 1 (10.0) |
| PSS-10 | 4 (33.3) | 5 (41.7) | 6 (50.0) | 4 (33.3) | 2 (16.7) | 3 (25.0) |

MCIDs: FACT-G = 3 points; FACIT-F = 3 points (McNeely et al., 2020); Balance = 24 seconds (Goldberg, Casby, & Wasielewski, 2011); SROM = >10°; MVPA = 26min/week (Hur et al., 2019); Sit to Stand = 2.6 repetitions (Wright, Cook, Baxter, Dockerty, & Abbott, 2011); PSS-10 = 2.66 points (Drachev et al., 2020)

*Missing Values: FACT-G = 1 (Non-HC); FACIT-F = 1 (Non-HC); Balance (R) = 2 (Non-HC); Balance (L) = 2 (Non-HC); SROM (R) = 2 (Non-HC); SROM (L) = 2 (Non-HC); MVPA = 1 (Non-HC); Sit to Stand = 3 (Non-HC); PSS-10 = 1 (Non-HC).
